# EXercise TheRApy in Mesothelioma feasibility study (EXTRA-Meso Feasibility) – Protocol of a randomised feasibility study

**DOI:** 10.1101/2025.08.25.25333696

**Authors:** S. Tsim, K.G. Blyth, J. Moore, A. Marcu, Z. Merchant, C. MacRae, M Evison

## Abstract

**Introduction:** Malignant Pleural Mesothelioma (MPM) is commonly associated with high symptom burden, cancer cachexia and poor health-related quality of life (HRQOL). Uptake of systemic anti-cancer treatment is low, often due to reduced physical fitness associated with a diagnosis of MPM and advanced age at diagnosis. There is evidence to support exercise therapy in other cancers, including lung cancer, but there is limited evidence for the role of exercise therapy in MPM. EXTRA-Meso feasibility study is a randomised feasibility trial of exercise therapy versus standard care in patients with a diagnosis of MPM.

**Methods and Analysis:** Patients with a diagnosis of mesothelioma, with performance status 0 – 2, clinical frailty score ≤5 will be randomised 1:1 between exercise intervention, where the patient is provided with a personalised exercise and wellbeing programme, after being assessed by a physiotherapist or qualified exercise professional with specific cancer training, or standard care (routine clinical follow-up). The primary objective is to determine whether sufficient numbers of patients, defined as 40 patients over 12 months, can be recruited and randomised to justify a subsequent phase III trial examining the role of exercise therapy in maintaining or improving HRQOL in patients with MPM. Recruitment will take place across 2 UK mesothelioma centres (Glasgow and Manchester). Secondary objectives include assessment of barriers to recruitment and retention, and the safety and tolerability of the intervention and study assessments.

**Ethics and Dissemination:** Protocol approved by West Midlands – South Birmingham Research Ethics Committee (Ref 23/WM/0186). Results will be presented at international scientific meetings and published in peer-reviewed journals.

**Trial registration:** ISRCTN24557328

## Introduction

Malignant Pleural Mesothelioma (MPM) is an invasive thoracic malignancy, strongly associated with asbestos exposure, which accounts for approximately 2500 deaths per annum in the United Kingdom. MPM is associated with a poor median overall survival of one year. High symptom burden, anxiety and depression are common in MPM, even in those with good performance status (PS). [1,2] Patients with MPM often report poor health-related quality of life (HRQOL), which is itself, associated with poorer survival. [3,4] The treatment of MPM typically involves systemic anti-cancer therapy (SACT), receipt of which may be associated with better maintenance of HRQOL compared with those who are managed with best supportive care. [5] However, SACT uptake in MPM is low, often due to reduced physical fitness and functional performance in patients who are frequently in their 7^th^ or 8^th^ decade of life, due to prolonged latency period between asbestos exposure and disease development. Maximising and maintaining HRQOL is a key goal for all MPM patients and maximising treatment opportunity has never been more important, following recent evidence supporting treatment for up to 2 years with immunotherapy in MPM. Combination nivolumab and ipilimumab significantly improves overall survival in unresectable MPM compared to standard chemotherapy in the first-line setting. [6] This survival benefit appears amplified in patients with non-epithelioid histology, with maintenance or improvement in HRQOL in responding patients. [7] This is particularly important in this sub-group, in whom standard chemotherapy is rarely effective and symptom burden is frequently high with poor HRQOL. [5]

### Exercise Therapy

Exercise therapy is a rational approach to improving HRQOL and could also maximise treatment opportunities in MPM. This reflects a growing body of evidence for exercise interventions in other cancers. In non-small cell lung cancer (NSCLC), pre-operative exercise therapy in patients undergoing resection is associated with improved cardio-respiratory fitness, reduced post-operative complications and improved HRQOL. [8] Olivier et al also reported that a home pulmonary rehabilitation programme was safe and feasible in patients with lung cancer and a small number of patients with MPM. [9] It has recently been reported that low skeletal muscle mass (sarcopenia), a feature of the cancer cachexia syndrome, is independently associated with adverse survival in MPM. This effect was more marked in thoracic musculature ipsilateral to the tumour [10] and was observed if sarcopenia was present prior to starting chemotherapy and if patients lost skeletal muscle mass during treatment. [10] These features suggesting a dynamic deconditioning effect, driven directly by MPM, which could be predicted and modified to improve outcomes. In these studies, the presence of sarcopenia was also notably associated with systemic inflammation. [11] A recent systematic review and meta-analysis has confirmed poorer progression-free survival and overall survival in patients with sarcopenia pre-immunotherapy in solid tumours including lung cancer and melanoma. [11] In MPM, malnutrition and ‘pre-sarcopenia’ has previously been reported in 38% and 54% of patients respectively, with lower activity levels in pre-sarcopenic patients. [12] Exercise intervention in patients with sarcopenia may improve muscle mass, muscle strength and physical performance. [13] There is also evidence that exercise may have a role in breaking the vicious cycle of cancer, inflammation and cachexia [14]. Furthermore, there is evidence that exercise improves quality of life and reduces anxiety in patients with other cancers. [15,16] The ‘Prehab4Cancer’ programme, which this study will be utilising, reported improvements in functional fitness and quality of life in patients with NSCLC. [17] However, there is no evidence regarding the potential benefits of a tailored exercise, wellbeing and nutritional support intervention in mesothelioma. This is the focus of the current feasibility study and a planned future phase 3 trial.

The aim of this study is to examine the feasibility of a randomised trial of exercise therapy in MPM. It will define likely rate of recruitment based on performance in two prominent mesothelioma centres and identify barriers to recruitment and retention. Outcomes from the study will help us refine the design of a future definitive, potentially practice changing, phase 3 trial, including the number of recruiting centres required, and the optimum duration and format of the exercise intervention.

## Methods and analysis

### Study Design and Setting

EXTRA-Meso feasibility is a randomised, prospective feasibility study, recruiting from two UK centres (Glasgow and Manchester) over a period of 12 months. Site selection was based on access to a regional or national mesothelioma Multi-Disciplinary Team (MDT) and diagnosis of more than 25 cases per year. Glasgow is the hub for the Scottish Mesothelioma Network, serving a West of Scotland population which diagnoses approximately 65 – 75 new MPM cases per year. Greater Manchester similarly diagnoses approximately 50 new MPM cases per year and the recruiting centre serves as the regional centre for MPM. The overall study design is summarised in Figure 1. The planned study start and end dates are 31^st^ January 2024 and 31^st^ October 2025 respectively.

**Figure 1.**
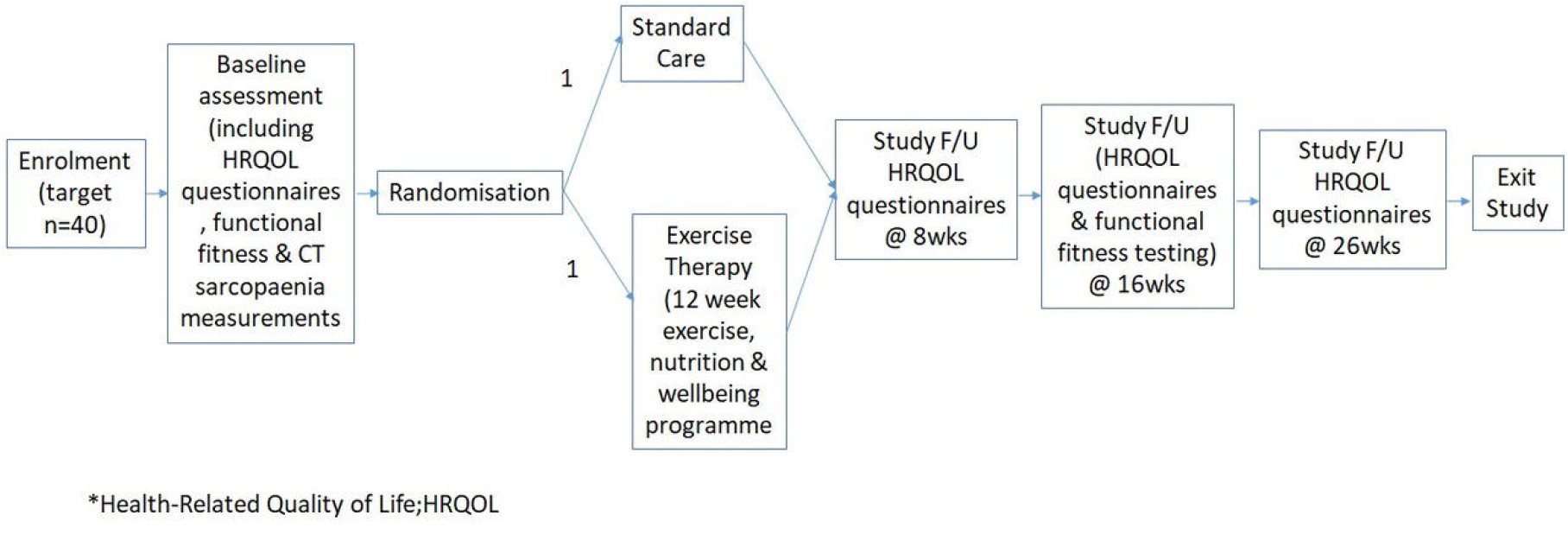
Study flowchart.

### Study Objectives and Outcome Measures

Study objectives and their associated outcome measures are summarised in Table 1.

**Table 1.** Primary and secondary objectives and their associated outcome measures.

| **Primary Objectives** | **Outcome Measures** |
| --- | --- |
| To determine the feasibility of recruiting patients with mesothelioma to an exercise therapy study | Number of patients recruited |
| **Secondary Objectives** | **Outcome Measures** |
| To determine barriers to study recruitment | Themes from qualitative interviews<br>Proportion of screened patients who are ineligible<br>Reasons for study ineligibility |
| To determine barriers to study retention | Themes from qualitative interviews<br>Study drop-out rate<br>Reasons for study drop-out |
| To determine safety and tolerability of the exercise intervention and study assessments | Intervention adherence rate<br>Adverse events<br>Health-Related Quality of Life questionnaire completion rate |

### Eligibility Assessment

All patients will be subject to the following eligibility criteria. Patients are eligible for the study if they meet all inclusion criteria and none of the exclusion criteria apply. All patients, regardless of treatment status are potentially eligible. The inclusion criteria are: diagnosis of MDT confirmed by mesothelioma MDT, Eastern Cooperative Oncology Group (ECOG) performance status (PS) 0 – 2, clinical frailty score (CFS) ≤5, informed written or remote consent. The exclusion criteria are: Performance Status ≥3, Clinical Frailty Score ≥6, patient unlikely to be able to participate in an exercise programme based on clinician or physiotherapist’s judgement. These PS and CFS constraints were considered clinically appropriate for this feasibility study, after consultation with our patient and public involvement group. Based on audit data from the Scottish Mesothelioma Network and the National Mesothelioma Audit [18], between 67 – 89% of patients are PS 0 – 2 at time of diagnosis with mesothelioma. These eligibility criteria should therefore encompass the majority of our patient population.

### Identification of participants and consent

Potential participants will be identified at mesothelioma MDTs, and at respiratory and oncology outpatient clinics. Patients will be assessed by the respiratory physician/site principal investigator co-ordinating their clinical care or delegated members of the research team. Potential participants will be given sufficient time to consider the participant information sheet, discuss with family or friends and the opportunity to ask questions about the study, before deciding whether they wish to participate, same day consent is permissible. Consent can be obtained face to face or remotely in order to maximise potential opportunities for consent, particularly as the recruiting centres offer regional mesothelioma services. Detailed screening logs will be kept to record reasons for non-enrolment. Should the patient read the patient information sheet but decline to take part, they may be asked to consent to a semi-structured interview with a member of the study team, to explore their reasons for declining to participate, to inform future study design. Potential participants who decline to participate will be signposted to local community activities and services that are available to promote long-term health improvement if they wish. There is no time limit on time of diagnosis to study enrolment. Eligibility will be confirmed by either a nurse or medical practitioner.

### Randomisation and study procedures

Following consent, baseline assessments will be completed by a member of the research team. A summary of the required data, screening tools, functional fitness assessments and HRQOL tools at each study visit is provided in Table 2. Patients whose Malnutrition Universal Screening Tool (MUST) score indicates malnourishment will be referred NHS dietetic services.

**Table 2.** Summary schedule of assessments.

| **Data/Assessments** | **Baseline** | **Week 8** | **Week 16** | **Week 26** |
| --- | --- | --- | --- | --- |
| **Patient demographics** | X |  |  |  |
| **Performance status** | X |  |  |  |
| **Mesothelioma date of diagnosis, subtype, planned/current systemic anti-cancer treatment (SACT)** | X |  |  |  |
| **Relevant clinical updates, e.g. treatment changes** |  | X | X | X |
| **Body Mass Index** | X |  |  |  |
| **MUST score** | X |  |  |  |
| **SARC-F score** | X |  |  |  |
| **Functional Fitness Assessments** |  |  |  |  |
| **Six Minute Walk Test (6MWT)** | X |  | X |  |
| **Hand Grip Strength** | X |  | X |  |
| **30 second sit-to-stand test** | X |  | X |  |
| **Health-Related Quality of Life Questionnaires** |  |  |  |  |
| **EORTC QLQ-C30** | X | X | X | X |
| **EQ-5D-5L** | X | X | X | X |
| **SF-36** | X | X | X | X |
| **HADS** | X | X | X | X |
| **sWEMWBS** | X | X | X | X |

\*Malnutrition Universal Screening Tool, MUST; Strength, Assistance with walking, Rised from a chair, Climb stairs, and Falls, SARC-F; European Organisation for Research and Treatment of Cancer, EORTC; Euroqol quality of life survey, EQ-5D-5L; 36-item Short Form survey, SF-36; Hospital and Anxiety Depression Scale, HADS; Short Warwick Edinburgh Mental Wellbeing Scale, sWEMWBS

### HRQOL tools

Validated HRQOL tools were selected to ensure a broad assessment of cancer-specific symptoms, physical, emotional and social functioning and mental health. The range of questionnaires will also allow comparison the results of HRQOL outcomes in the future phase III study with other malignant and non-malignant populations. If there is a high study drop-out due to questionnaire burden during the study, these will be reviewed. If required, questionnaires can be posted out to patients and returned to the study team using a pre-paid addressed envelope. Study team members can assist participants with completion of these questionnaires remotely if required. This was designed to be pragmatic and minimise research study visit burden for patients.

### Body composition measurements

Routinely-acquired clinical computed tomography (CT) scans, if available, will be used to determine whether body composition measurements (skeletal muscle area and visceral fat index) can be reliably measured using clinical CT scans, given the variability in image acquisition that can occur. Images will be analysed using CoreSlicer [19], a validated web-based application which enables semi-automated segmentation of muscle and fat. The ability to reliably record body composition measurements using clinical CT scans at baseline in particular is of interest, as we plan to assess in a future phase III study, should this study prove feasible, whether these measurements serve useful as predictive markers for subsequent response to exercise therapy. Within this study, measurements will not be used to compare differences longitudinally or between intervention groups.

### Randomisation

On completion of baseline assessments, patients will be randomised on 1:1 ratio using random permuted blocks, to receive exercise intervention or standard care using a validated online system (www.sealedenvelope.com). Within this feasibility study, there will be no minimisation criteria, e.g. by histological subtype or treatment type (chemotherapy versus immunotherapy versus none), due to participant numbers.

### Post-randomisation standard care arm

Participants randomised to Standard Care will undergo routine clinical follow-up, with study assessments completed as detailed in Table 2.

During the follow-up period, patients will be sign-posted to local community activities and services that are available to promote long-term health improvement. It will be made clear to participants within the standard care arm that they are free to undertake exercise or increased physical activity if they wish. Any increase in physical activity or participation in local generic community exercise programmes will be recorded at follow-up visits.

### Exercise therapy (intervention) arm

Participants randomised to the Exercise Therapy arm will undergo an individualised assessment by either a physiotherapist or a qualified exercise professional with specialist cancer training, either in hospital or in the community within 7 days of randomisation. Following this initial assessment, patients will receive a personalised exercise, wellbeing and nutritional support package. The exercise programme will incorporate 3 sessions per week. One session per week will be supported by the physiotherapist or qualified exercise professional and the other 2 sessions will be independent sessions done in the participant’s own home or local health club/gym if available. The total duration of the programme is 12 weeks. The exercise programme will incorporate both cardiovascular and resistance training and will be progressive throughout the duration of the programme. The exercise programme will be tailored to take into account the participant’s current level of fitness and familiarity/confidence with exercise, available equipment and injuries. This detailed assessment ensures that the exercise programme is always set at a level appropriate to each individual patient and is realistic and achievable. If patient’s functional status does decline during the course of their study participation, the exercise programme can be altered to accommodate this by the physiotherapist or exercise specialist, if the patient wishes to continue within the study. Participants will be given a diary to record completion of each exercise session, in order to help determine the acceptability of the exercise intervention. Activity trackers will not be used due to budget constraints. Follow-up visits will include the assessments detailed in Table 2.

### Sample Size and Statistical Considerations

The current feasibility study will aim to recruit 40 patients. No formal sample size is feasible or appropriate. Combined, the recruiting centres involved in this study diagnose approximately 115 MPM per year, of these we estimate approximately 70% (n=80) will be eligible, given the relatively broad eligibility criteria. Previous randomised intervention trials in mesothelioma have reported the proportion of eligible patients who subsequently consent is approximately 50%. [20] We therefore estimated that the 2 recruiting centres could recruit the target number of patients over 12 months. 40 patients should be sufficient to allow identification of barriers to recruitment and retention (the latter informing the likely drop-out rate for the future phase 3 trial) and the duration and design of the exercise therapy intervention.

The primary objective of the future phase 3 trial will be to determine whether exercise therapy results in a clinically meaningful improvement in HRQOL at 16 weeks post-randomisation. Koller et al reported a minimum important difference in quality of life (using the validated European Organisation for Research and Treatment of Cancer (EORTC) Core Questionnaire (QLQ C30) of +/-5 to 10 based on 2 randomised trials in NSCLC and MPM. [21] Assuming a mean population score of 60 (SD 20) based on QoL scores previously reported in other mesothelioma studies [1,2,4,5], we currently estimate that a sample size of 84 patients in each arm is required to detect a 10-point improvement in EORTC QLQ C30 quality of life scores, with 90% power at 5% two-sided significance level. With a currently assumed 25% drop-out rate, based on the ‘Prehab4Cancer’ dropout rate in patients with NSCLC [17], we estimate that we would require a total sample size of 224 patients. Data from the current feasibility study will (a) provide a more robust estimate of baseline mean HRQOL score (based on the population score recorded from all patients at baseline assessment), (b) allow a more precise estimate of drop-out rate based on an optimised exercise intervention and (c) allow us to calculate the number of centres needed based on the recruitment rate achieved.

### Statistical analysis plan

The primary outcome measure of the number of patients recruited will be expressed as a mean monthly rate over the complete trial period. Recruitment at each centre, reasons for study ineligibility, rate of study drop-out, reasons for study drop-out, HRQOL questionnaire completion and intervention adherence rate will be reported by simple descriptive statistics or proportions where appropriate. Differences in HRQOL scores by mode of delivery (postal versus face-to-face) will be assessed using mixed effect models with random intercepts. Missing data will be handled using mean or multiple imputation depending on the randomness of the missing data. Intra-class correlation co-efficient will be used to assess inter-observer agreement for body composition measurements.

### Feasibility Success Metrics

Feasibility success metrics, based on recommendations by Lewis et al. [22] collected during this study will determine if it is feasible to proceed to a phase 3 randomised trial and defined as:

- Green (proceed with no/minor changes to study design)
- Amber (significant changes to study design before proceeding)
- Red (do not proceed)

Recruitment

- Green = ≥30 patients (≥75% target recruitment)
- Amber = 23 – 29 patients
- Red = ≤22 patients (≤55% target recruitment)

### Qualitative Interviews

We will aim to conduct 5 x semi-structured interviews with eligible patients who decline the study to explore potential barriers to recruitment. A further 10 – 20 study participants and their carers (including patients who drop-out of the study before completion and those who complete the study) will be invited to a semi-structured interview to review their views on barriers to study retention, outcome measures, design of the phase 3 study and optimum duration and format of the exercise intervention. Participants randomised to intervention will be asked whether they received all the components of the interventions, and whether they completed 3 sessions of exercise per week as intended. The psychological and social barriers to participation will be explored in the qualitative interviews. The target sample size for interviews is based on recommendations for qualitative research in feasibility studies by O’Cathain et al. [23] Thematic saturation is not anticipated given the likely low participant numbers. Interviews will be conducted remotely or in person, depending on patient wishes. Audio transcriptions of the interviews will be analysed using a five-step framework analysis [24]: (1) familiarisation with the data; (2) identifying a thematic framework; (3) indexing; (4) charting; and (5) mapping and interpretation. The analysis will be mostly deductive as it will specifically focus on facilitators and barriers to study retention and to the uptake and experiences of the exercise intervention. The interviews will form the qualitative process evaluation of the intervention and provide insight into the mechanisms of impact of the intervention. All interviews will be analysed and coded by a single researcher (AM). Analysis will be informed by the Theoretical Framework of Acceptability of healthcare interventions. [25–27]

### Patient and Public Involvement

Patients had direct input on the design of this study after meeting with several patients with mesothelioma to ascertain their views on eligibility criteria, randomisation, study interventions and outcome measures. The patients interviewed included those had current or previous treatment with chemotherapy or immunotherapy, and patients who were treatment naïve, and were all of varying performance status.

### Definition of end of study

The end of study definition will be the date of last data capture, which will be met when all outstanding data has been returned from all sites, all required data queries have been resolved and the database is finalised for analysis.

### Safety considerations

All adverse events (AEs) and serious AEs (SAEs) thought to be related to study procedures will collected during routine clinical and study follow-up visits. All AEs and SAEs will be recorded in the patient’s medical records and reported to the trial sponsor.

### Ethics and dissemination

This study was approved by the West Midlands – South Birmingham Research Ethics Committee (REC reference 23/WM/0186). Patients will have the right to withdraw from the study at any time, without giving a reason, and this will not affect their clinical care or legal rights. Data already collected with consent will be retained and used in the study, unless the patient specifically withdraws their consent for its use. EXTRA-Meso feasibility results will be presented at scientific meetings and published in peer-reviewed journals.

## Data Availability

All data produced in the present study are available upon reasonable request to the authors

## Authors’ contributions

ST, KGB, JM, AM, ZM, CM and ME contributed to the conception and/or design of the work. KGB, JM, AM, ZM, CM and ME were involved in revising the work critically for important intellectual content, final approval of the version to be published, and agree to be accountable for all aspects of the work in ensuring that questions related to the accuracy or integrity of any part of the work are appropriately investigated and resolved. ST provided principal contribution to the conception and design of the work; drafting the work; final approval of the version to be published; and agrees to be accountable for all aspects of the work in ensuring that questions related to the accuracy or integrity of any part of the work are appropriately investigated and resolved. ST is the guarantor.

